# Prevalence and incidence of stress, depression, and anxiety symptoms among Brazilians in quarantine across the early phases of the COVID-19 crisis

**DOI:** 10.1101/2021.09.07.21263246

**Authors:** Miguel Blacutt, Alberto Filgueiras, Matthew Stults-Kolehmainen

## Abstract

**Objective:** The present study aimed to measure the prevalence and incidence of stress, depression, and anxiety symptoms in Brazilians during the COVID-19 pandemic.

**Method:** We assessed 103 (54 women, 49 men) participants online in three periods of the pandemic: March 2020 (T1), April 2020 (T2), and June 2020 (T3). Prevalence and incidence were identified when mental health scores were two standard deviations above the mean compared to normative data. Mental health indicators were measured using the Perceived Stress Scale, the Filgueiras Depression Index, and the State-Trait Anxiety Inventory – State Subscale.

**Results:** At T1, 89% of individuals were below cut-off scores for stress, anxiety, and depression, which dropped to 35% by T3. Stress prevalence was 1.9% at T1, 7.8% at T2, and 28.2% at T3. Depression prevalence was 0% at T1, 23.3% at T2, and 25.2% at T3. State anxiety prevalence was 10.7% at T1, 11.7% at T2, and 45.6% at T3. Stress incidence increased by 7.8% from T1 to T2, and 23.3% from T2 to T3. Depression incidence increased by 23.3% from T1 to T2, and 15.5% from T2 to T3. Anxiety incidence increased by 9.7% from T1 to T2, and 39.8% from T2 to T3. Stress severity scores significantly increased from 16.1±8.7 at T1 to 23.5±8.4 at T2, and 30.3±6.0 at T3. Depression severity scores significantly increased from 48.5±20.5 at T1 to 64.7±30.2 at T2, and 75.9±26.1 at T3. Anxiety increased from 49.0±13.4 at T1 to 53.5±12.5 at T2 and 62.3±13.4 at T3. Females had significantly higher anxiety scores than males by T3 (66.7±11.8 vs. 57.4±13.5).

**Conclusion:** Prevalence and incidence of stress, depression, and anxiety significantly increased throughout the pandemic. The largest increase in stress and anxiety occurred between T2 and T3, and between T1 and T2 for depression. Severity of stress, depression, and anxiety increased throughout the study.

## Introduction

The COVID-19 pandemic began to escalate in Brazil in mid-March, 2020 with the confirmation of the first cases of community transmission (Brazil Ministry of Health, 2021). Researchers have been attempting to understand the mental health consequences of the COVID-19 crisis, as well as the environmental and behavioral factors that might be linked to psychological distress or other potential disorders. However, there is a lack of epidemiologic data for mental illness in Brazil in response to the emergence of COVID-19 (Filgueiras and Stults-Kolehmainen, 2021; Goularte et al., 2021; Passos et al., 2020; Filgueiras and Stults-Kolehmainen, 2020). Understanding the prevalence and incidence of stress, depression, and anxiety symptoms among people in quarantine due to COVID-19 is pivotal as it sets the stage for future interventions and to aid individuals seeking help from mental health professionals as a result of the ongoing pandemic.

Previous studies aimed to understand the relationship between various behavioral and psychosocial factors with psychological distress, depression, and anxiety symptoms (Filgueiras and Stults-Kolehmainen, 2021; Goularte et al., 2021) during the COVID-19 pandemic. The results suggested that individuals of female gender are more likely to show higher severity of psychiatric symptoms. Behaviors such as engaging in exercise and physical activity, eating healthy and engaging in tele-psychotherapy are associated with less mental health problems during quarantine. Those participants who reported the need to leave their homes to go to work, regardless of the nature of their jobs, showed more severe symptoms (Abreu et al., 2020; Filgueiras and Stults-Kolehmainen, 2021). Social isolation during quarantine has played a role in the development of mental illness, particularly for depression compared to anxiety (Passos et al., 2020). Additionally, low income, lower education and self-reported history of previous psychiatric illness were strongly associated with higher severity of symptoms (Goularte et al., 2021)

Broadly, prevalence can be defined as the proportion of people who have a condition at a given time point while incidence refers to the proportion of people who develop a condition throughout a period of time (Porta, 2008). The prevalence of symptoms of depression and anxiety has been examined during the first three months of quarantine due to COVID-19 (Goularte et al., 2021; Passos et al., 2020). At least one symptom of anxiety was found in 81.9% of a sample of 1,996 Brazilians, whereas depression was reported by 68% of these volunteers (Goularte et al., 2021). Similar results were found in a Brazilian and Portuguese study with samples from the two countries. At least one symptom of anxiety was reported by 71.3% of the participants (mild anxiety was present in 43.1%), 24.8% declared at least one depression symptom and 23.8% presented both depression and anxiety. Results also showed significant prevalence of other psychiatric symptoms: anger (64.5%), somatic symptoms (62.6%) and sleep problems (55.3%). Additionally, post-traumatic stress disorder (PTSD) symptoms were present in 65.8% of the sample (Goularte et al., 2021). Those findings are similar to previous results with other pandemics in different countries, such as the Middle-East Respiratory Syndrome (MERS) (Lee et al., 2018) and the Severe Acute Respiratory Syndrome (SARS) (Wu et al., 2005). Other researchers who investigated mental illness during the COVID-19 pandemic in other countries also found an increased prevalence, such as the U.S.A. (Liu et al., 2020), China (Huang and Zhao, 2020), India (Rehman et al., 2021), Germany (Bäuerle et al., 2020), Italy (Rossi et al., 2020), Turkey (Özdin and Bayrak Özdin, 2020) and Bangladesh (Islam et al., 2020). A variety of psychosocial factors may account for the pattern of results found for prevalence of mental health problems. For example, Lee et al. (2018) found that the risk of medical staff reporting PTSD symptoms during the MERS epidemic was higher due to exposure to contamination and their perception of personal health risk. Wu et al. (2005) reported similar findings during the SARS epidemic; they presented results pointing out that PTSD, depression, and anxiety symptoms were associated with perception of risk.

Filgueiras and Stults-Kolehmainen (2021) investigated the prevalence of severe depression, anxiety, and psychological stress among 360 Brazilian volunteers between March and April of 2020. They adopted the criteria of two standard deviations above the mean (2 SD) from the normative data of validated psychometric instruments. Results showed that 9.7% of the sample showed psychological distress above the clinical cut-off criteria, while 8.0% and 9.4% of participants were also above for depression and anxiety, respectively. The World Health Organization collected prevalence data in Brazil (World Health Organization, 2017) three years before the quarantine and found a lower prevalence of depression (5.8%). Nonetheless, anxiety was similar (9.3%), perhaps indicating some stability during quarantine. Importantly, data from these previous COVID-19 studies were gathered between March and May.

This study aimed to identify prevalence and incidence of severe psychological stress, depression, and anxiety among Brazilians during the COVID-19 pandemic. We chose to include distress, depression and anxiety because these are the are common psychological symptoms observed in other epidemics of acute respiratory syndromes (Brown et al., 2020; Lee et al., 2018; Wu et al., 2005) and during COVID-19 (Bäuerle et al., 2020; Filgueiras and Stults-Kolehmainen, 2021; Goularte et al., 2021; Huang and Zhao, 2020; Islam et al., 2020; Liu et al., 2020; Özdin and Bayrak Özdin, 2020; Passos et al., 2020; Rehman et al., 2021; Rossi et al., 2020). We hypothesize that the prevalence and incidence of stress, depression, and anxiety throughout the COVID-19 pandemic will increase at each time point, and that scores for each of these conditions will also increase across time. Furthermore, we hypothesize that anxiety scores would increase higher in females compared to males. We built our hypothesis on previous evidence that suggests that stressful events combined with prevention measures against COVID-19 contamination, such as social isolation, quarantine, and confinement, may be associated with increased prevalence of mental illness (Brown et al., 2020; Liu et al., 2020). Additionally, our hypothesis is informed by previous literature showing higher health-anxiety and occurrence of anxiety disorder in females compared to males (Wang et al., 2021; Stults-Kolehmainen et al., 2014). Information about prevalence and incidence may support decision making about strategies and policies to address mental illness in places where the COVID-19 pandemic is still a problem.

## Methods

### Participants

This present longitudinal study assessed mental health outcomes in volunteers (*N*=103) during the initial stages of the COVID-19 pandemic in Brazil. We collected data over three periods of the COVID-19 pandemic curve (Pinto et al., 2020): Time 1, the first cases of community transmission (T1, March 20, 2020 to March 25, 2020); Time 2, the acceleration of the curve (T2, April 15, 2020 to April 20, 2020); and Time 3, the continued acceleration of the curve (T3, June 25, 2020 to June 30, 2020). Inclusion criteria were being a Brazilian or a foreigner living in Brazil during the COVID-19 pandemic who adhered to quarantine measures. Exclusion criteria were being under the age of 18 years old (*N*=1) and being a diagnosed psychiatric patient currently under treatment (*N*=6), since those participants are already above clinical cut-off criteria and could show comorbid mental illnesses which would inflate prevalence numbers. Volunteers who reported being of non-binary gender (*N*=9) were also excluded due to the low number of participants, which impaired statistical analyses; nonetheless, they were considered in this paper’s discussion. Respondents were asked to complete a 45-minute online Google Forms questionnaire regarding demographic information and COVID-19-related mental health outcomes. The informed consent document was presented before the questionnaires, and the consent was a requirement for participation. The Ethics Committee at Rio de Janeiro State University approved all procedures (report #2.990.087).

### Measures

We adopted three validated and normalized measures to ensure good quality of data; one instrument for each psychological dimension: psychological stress, depression, and anxiety. Severe stress, depression, and anxiety were identified if the respective mental health score was two standard deviations above the mean compared to normative data. Demographic information was collected through a simple questionnaire containing: gender (male, female and non-specific), age (in years) and risk for COVID-19 (“Do you have any current disease that increases your risk for COVID-19 lethality?”—yes/no). All instruments were presented in Brazilian Portuguese.

#### Perceived Stress Scale (PSS-10)

The PSS-10 (Cohen and Williamson, 1998) is a 10-item measure that assesses the state of perceived stress in the last month. Participants respond to questions on a 5-point Likert-type scale that ranges from “0-never” to “4-very often” in regard to the frequency of those feelings or behaviors. Examples of questions are: “In the last month, how often have you been able to control irritations in your life?” and “In the last month, how often have you felt that you were unable to control the important things in your life?” Items 4, 5, 7 and 8 are reverse scored before summing to develop a single total score. Norms were developed for men and women separately (Cacciari et al., 2016), where the average for men was 20.7 (SD = 6.9) and the average for women was 23.2 (SD = 5.8). Scores above 34.5 and 34.8 for men and women, respectively, were considered to indicate severe stress.

#### Filgueiras Depression Inventory (FDI)

The FDI (Filgueiras et al., 2014) is a 20-item inventory of words that are related to depression-like symptoms according to the DSM-V. Respondents link each of these words to their own feelings in the last fortnight. A Likert-type scale response set contains six categories of endorsement ranging from “0-not related to me at all” to “5-totally related to me”. Examples of items are “sadness”, “death”, “displeasure” and “guilty”. The total score is the sum of all items. The reference mean is 53.3 (SD = 22.6) with 99.1 or higher indicating a clinical cut-off point (2 SD) for depressive symptomology for both males and females.

#### State and Trait Anxiety Inventory—State subscale (STAI-S)

The STAI-S is one in a set of two subscales developed to assess trait and state anxiety in adults (Spielberger et al., 1983). Trait anxiety refers to stable characteristics of an individual that facilitate the occurrence of anxiety-like symptoms and behaviors. On the other hand, state anxiety comprises how one feels in the moment the inventory is completed. The state anxiety subscale has a 20-item structure that is answered with a 4-category Likert scale. The STAI-S responses range from “1-not at all” to “4-very much so”. Examples of items are “I feel calm”, “I feel nervous” and “I am presently worrying over possible misfortunes”. Items 1, 2, 5, 8, 10, 11, 15, 16, 19 and 20 are reverse scored before summing responses to provide a total score. Gender-specific reference means are 36.5 (SD = 14.3) for male respondents and 43.7 (SD = 12.7) for female respondents, with clinical cut-offs (> 2 SD) for severe acute anxiety being above 65 for men and above 69 for women (Pasquali et al., 1994)

### Data analysis

All variables were normally distributed with skewness and kurtosis statistics within the range of -2.0 and 2.0. Prevalence was calculated using the percentage of participants above cut-off points in each period (T1, T2, and T3). Incidence of stress, depression, and anxiety was indicated by the percentage of participants whose scores were below cut-offs in the previous period data collection but were above these cut-offs in the next. We estimated three incidence percentages: (i) between times 1 and 2, (ii) 2 and 3 and (iii) 1 and 3. 95% confidence intervals were estimated as described by Altman et al. (2000). Furthermore, differences in PSS-10, FDI, and STATI-S between data-collection (time), gender, perception of risk (binary variable indicating whether individual indicated having a pre-existing comorbidity perceived to increase risk of COVID-19 fatality) and their interaction were evaluated using linear mixed effects models, where a random intercept was generated for each participant. Tukey post-hoc tests were used to find pairwise differences in main effects and interactions. Pearson correlations were performed to investigate linear associations between all mental health outcomes and age at each data collection point. Our results and interpretations were based on a significance set at *p* < .05. All analyses were performed using R Statistical Software (Version 4.02; R Foundation for Statistical Computing, 498 Vienna, AT).

## Results

Table 1 shows characteristics of our participants and descriptive data on mental health outcomes. This longitudinal study included 103 participants (32.3±10.7 years; 52% female). 44 volunteers (42.7%) reported no chronic illness that increased lethality of COVID-19 due to comorbidities at time 1 (64% female, 36% male). Of the 59 individuals who reported having higher risk for lethality for COVID-19, 26 (44%) were female and 33 (56%) were male.

**Table 1.** Participant age, stress, depression, and anxiety scores stratified by time point, gender, and perception of higher risk for COVID-19 lethality resulting from pre-existing illness

|  | Age | Stress (PSS-10) |  |  | Depression (FDI) |  |  | Anxiety (STATI-S) |  |  |
| --- | --- | --- | --- | --- | --- | --- | --- | --- | --- | --- |
|  | Time | Time | Time | Time | Time | Time | Time | Time | Time | Time |
|  | 1 | 1 | 2 | 3 | 1 | 2 | 3 | 1 | 2 | 3 |
| **Overall** |  |  |  |  |  |  |  |  |  |  |
| Total | 32.3 | 16.1 | 23.5 | 30.3 | 48.5 | 64.7 | 75.9 | 49.0 | 53.3 | 62.3 |
| (N=103) | (10.8) | (8.7) | (8.4) | (6.0) | (20.5) | (30.2) | (26.1) | (13.4) | (12.4) | (13.4) |
| **Gender** |  |  |  |  |  |  |  |  |  |  |
| Female | 31.9 | 16.9 | 23.9 | 31.6 | 50.5 | 65.9 | 81.7 | 50.3 | 53.7 | 66.7 |
| (N=49) | (11.1) | (8.5) | (23.2) | (28.9) | (19.2) | (27.1) | (30.0) | (12.7) | (11.2) | (11.8) |
| Male | 32.7 | 15.2 | 23.2 | 28.9 | 46.2 | 63.5 | 69.5 | 47.6 | 52.8 | 57.4 |
| (N=54) | (10.5) | (8.9) | (9.2) | (6.4) | (21.8) | (33.5) | (22.5) | (14.1) | (13.7) | (13.5) |
| **Risk** |  |  |  |  |  |  |  |  |  |  |
| No | 30.1 | 17.6 | 23.6 | 31.2 | 51.0 | 69.4 | 84.0 | 51.6 | 54.0 | 68.8 |
| (N = 44) | (11.2) | (8.9) | (8.7) | (5.9) | (21.1) | (29.3) | (24.6) | (13.8) | (12.2) | (8.6) |
| Yes | 33.3 | 15.0 | 23.5 | 29.6 | 49.6 | 61.3 | 69.9 | 47.1 | 52.7 | 57.4 |
| (N = 59) | (10.4) | (8.4) | (8.2) | (6.1) | (20.0) | (30.6) | (25.8) | (12.9) | (12.6) | (14.5) |

The prevalence of severe stress symptoms was 1.9% (95% CI [0.5, 6.8]) at T1, 7.8% (95% CI [4.0, 14.6]) at T2 and 28.2% (95% CI [20.4, 37.5]) at T3. The prevalence of severe depression symptoms was 0% (95% CI [0, 3.6]) at T1, 23.3% (95% CI [16.2, 32.3]) at T2 and 25.2% (95% CI [17.8, 34.4]) at T3. The prevalence of severe anxiety-state symptoms was 10.7% (95% CI [6.1, 18.1]) at T1, 11.7% (95% CI [6.8, 19.3]) at T2 and 45.6% (95% CI [36.3, 55.2]) at T3. We observed greater prevalence of mental illness across the COVID-19 pandemic, as well as overlap between stress, depression, and anxiety. The progression of mental illness and the overlap between the conditions can be seen in Figure 1. Of note, there was no prevalence of severe depression, anxiety, or stress in 89% of individuals at T1, which dropped to 35% by T3.

**Figure 1.**
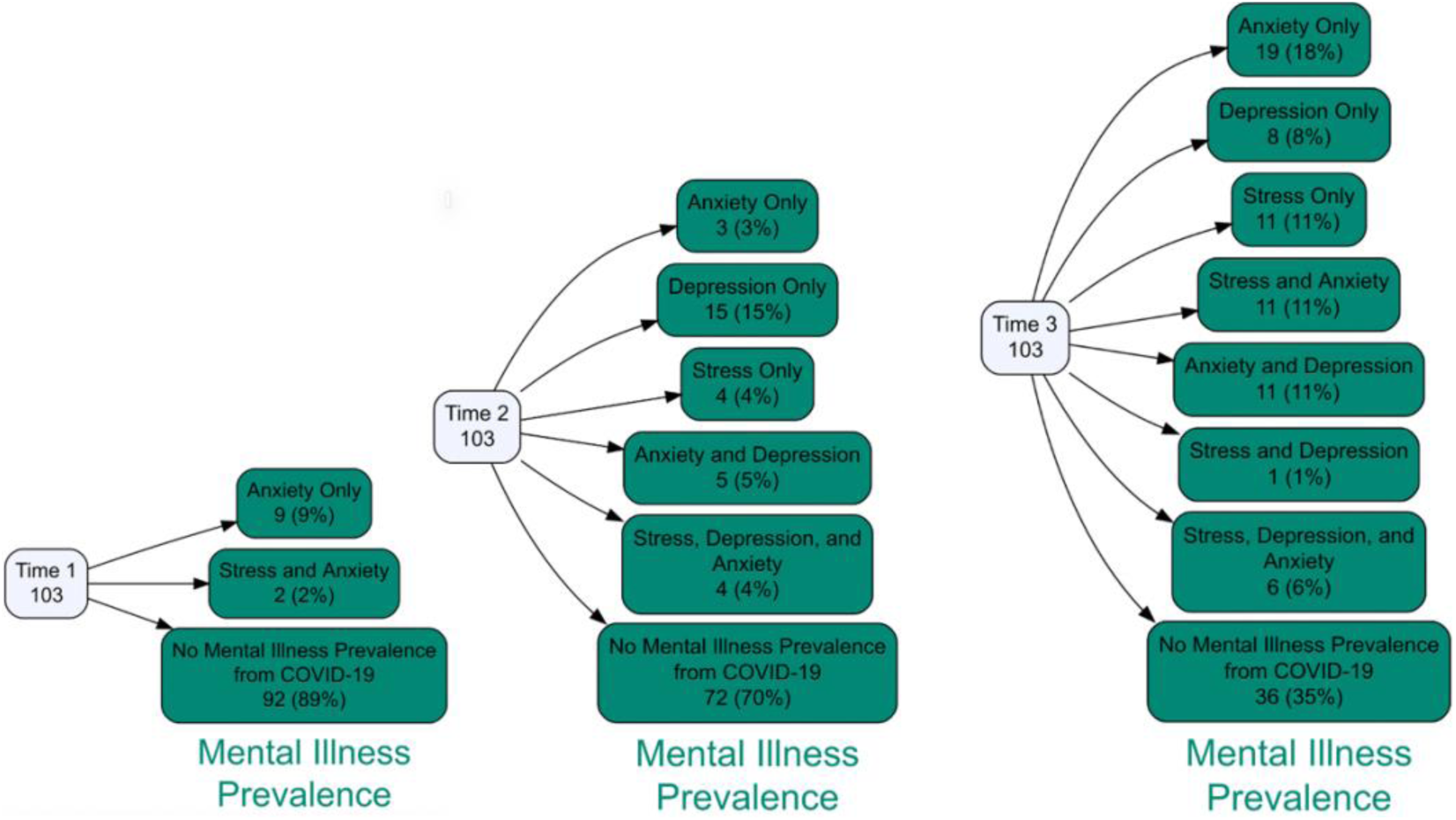
Prevalence of anxiety-alone, depression-alone, stress-alone, anxiety and depression, anxiety and stress, depression, and stress, all three, and neither at each time point of the COVID-19 pandemic

The results of the linear-mixed effects models for stress, anxiety, and depression are displayed in Figure 2. A main effect of time was found for PSS-10 (*F*(2, 198) = 103.5, p <.0001), however, there was no main effect of gender or risk on PSS-10 as well as no interactions. Tukey post-hoc tests revealed PSS-10 to be significantly higher at T3 compared to T2 (*t*(198) = -7.13, *p* < .0001) and T1 (*t*(198) = -14.4, *p* < .0001), while T2 was found to be significantly higher than T1 (*t*(198) = -7.23, *p* <.0001). A main effect of time was found for FDI (*F*(2, 198) = 52.9, *p* < .0001) however, there was no effect of gender or risk perception on PSS-10 in addition no interactions. FDI was found to be significantly higher at T3 compared to T2 (*t*(198) = -7.43, *p* = .0001) and T1 (*t*(198) = -10.3, *p* < .0001). Moreover, T2 was found to be significantly higher than T1 (*t*(198) = - 6.02, *p* <.0001). A main effect of time (*F*(2, 198) = 55.0, *p* < .0001) and risk perception (*F*(1, 99) = 5.55, *p* = .021) was found for STAI-S. Tukey post-hoc comparisons revealed that STAI-S was significantly higher at T3 compared to T2 (*t*(198) = -7.34, *p* < .0001) and T1 (*t*(198) = -10.2, *p* < .0001), while T2 was found to be significantly higher than T1 (*t*(198) = -2.82, *p* = .015). Furthermore, there was a significant difference between risk perception levels where individuals without a comorbidity had higher STAI-S scores than individuals with a comorbidity (*t*(99) = 2.36, *p* < .02), as is seen in Figure 3. A time-by-risk interaction was found for STAI-S scores (*F*(2,198) = 5.96, p = .003), and a Tukey post-hoc comparison showed that individuals without a comorbidity had higher STAI-S scores than those with a comorbidity (68.8±8.6 vs. 57.4±14.4; *t*(209) = 3.81, *p* = .003). Finally, a time-by-gender interaction was found for anxiety (*F*(2,198) = 3.17, p = .044), and a Tukey post-hoc comparison showed that females had higher anxiety scores than males at T3 (66.7±11.8 vs. 57.4±13.5; *t*(209) = 2.94, *p* = .04).

**Figure 2.**
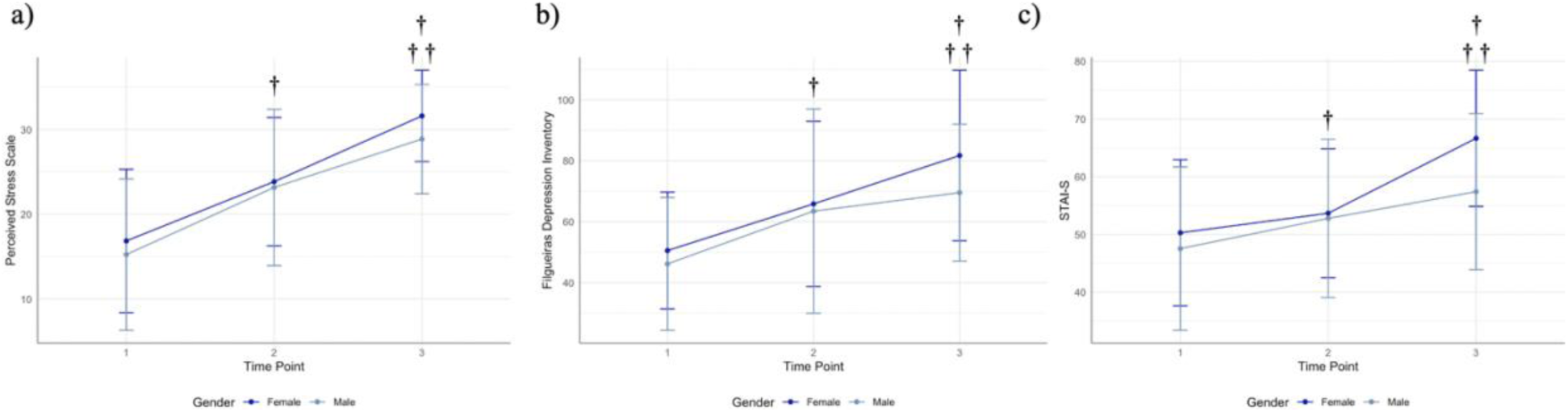
Stress, depression, and anxiety scores across each time point and stratified by gender. † indicates statistical difference from T1 (p < .05). †† indicates statistical difference from T2 (p < .05)

**Figure 3.**
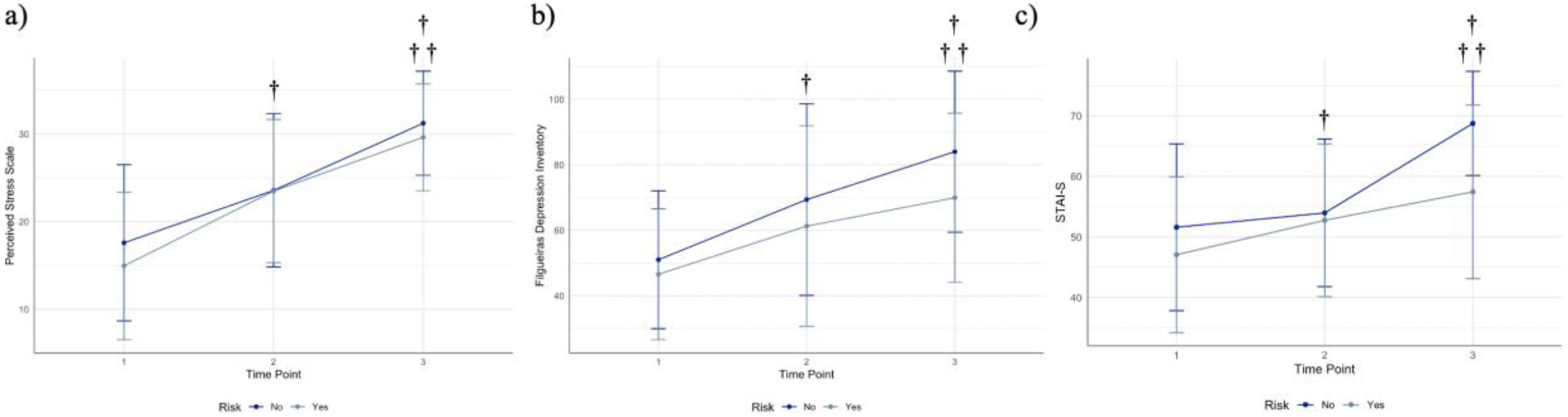
Stress, depression, and anxiety scores across each time point and stratified by binary perception of risk. † indicates statistical difference from T1 (p < .05). †† indicates statistical difference from T2 (p < .05)

Figure 4 depicts the correlation matrices between age and mental health outcomes at each data collection point. At T1, all correlations were statistically significant (*p*≤.05) and ranged from *r* = -.21 to *r* = .91. Age was significantly and inversely correlated with all mental health outcomes. Stress was strongly and positively correlated to both depression (*r* = .83) and anxiety (*r* = .91). Furthermore, there was a strong relationship between depression and anxiety (r = .82). At T2, age was only significantly associated with depression (*r* = -.45, *p* < .05). Moreover, only stress and anxiety were associated, though the relationship was strong (*r* = .88, *p* < .05). At the T3, there were no significant associations between any mental health outcomes or age.

**Figure 4.**
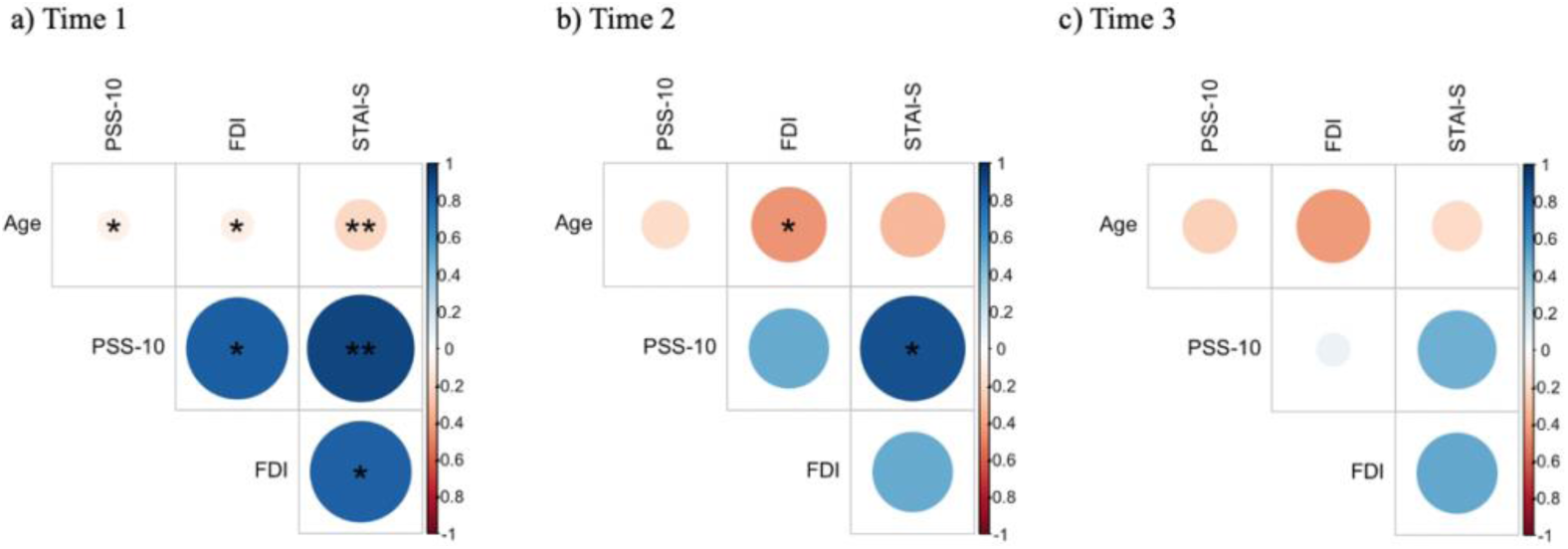
Pearson correlation matrix depicting relationship between age, stress, anxiety, and depression at each time point. Correlations conducted for each time point separately, a) T1, b) T2, and c), T3. * denotes *p* < .05, ** denotes *p* < .01, *** denotes *p* < .001. Darkness, color, and size of circle depict strength of correlation from *r* = -1 to 1.

## Discussion

Our findings highlight the increase of psychological symptoms throughout the early phase COVID-19 pandemic based on both prevalence and incidence. Prevalence of severe stress symptoms increased from 1.9% (T1) to 28.2% (T3). The same phenomenon occurred with depression, increasing from 0.0% % (T1) to 25.2 % (T3), and with anxiety, increasing from 10.7% (T1) to 45.6% (T3). Furthermore, our results show that the largest increase in incidence of stress and anxiety occurred between T2 and T3 (23.3% and 39.8%, respectively), while the largest increase in depression incidence occurred between T1 and T2 (15.5%). In 2017, the World Health Organization gathered worldwide data regarding mental health and found a prevalence of 5.8% for depression and 9.3% for anxiety among Brazilians (World Health Organization, 2017). Our results suggest that this sample had a lower prevalence of depression at the start of the COVID-19 pandemic, compared to the WHO’s data. Nevertheless, the acceleration phases of the epidemic curve showed higher depression prevalence compared to the WHO, by a magnitude of 17.5% and 19.4% at T2 and T3, respectively. Previous literature shows that depressive individuals during the COVID-19 pandemic tend to present melancholic, social isolation, grief and denying symptoms (Goularte et al., 2021; Huang and Zhao, 2020; Liu et al., 2020). On the other hand, regarding anxiety, time 1 of our data collection presented higher levels of prevalence by a magnitude of 1.4% when compared with those results from WHO. Furthermore, the second measurement exceeded the WHO’s previous data by a magnitude of 2.4% while the third measurement exceeded the WHO’s previous data by a magnitude of 36.3%.

The incidence of stress, depression, and anxiety among people in quarantine is coherent with other studies. Brown et al. (2020) reviewed previous data regarding mental health during other respiratory syndrome epidemics and concluded that stress, anxiety, anger, and depression are the most common psychiatric symptoms, and they tend to be more severe during quarantine. Our results corroborate with previous findings (Brown et al., 2020; World Health Organization, 2017) and add new information regarding the time period of quarantine that people may be in greatest need of help. Symptoms of stress, depression, and anxiety increased from the start of the pandemic to the continued acceleration phase recorded at time 3. Nonetheless, depression symptoms behaved differently; it began with a smaller prevalence compared to normative data (5.8%), then rose to 23.3% at time 2 and 25.2% at time 3.

Our analysis showed that gender had a significant interaction with time for anxiety, where females had higher anxiety scores than males by the third collection point (66.7±11.8 vs. 57.4±13.5). These results are consistent with Özdin and Bayrak Özdin (2020), who found that females had significantly higher anxiety and health anxiety, compared to male counterparts. Furthermore, Wang et al. (2021) found that females had 3.01 times the anxiety risk than males during the COVID-19 pandemic. These findings differ slightly from ours, as we found no significant difference in depression between genders. We speculate that these between-sex differences are explained by a higher likelihood for health anxiety in women compared to men, a higher incidence of lifetime anxiety disorder in women compared to men, and a larger proportion of healthcare workers being women (Xiong et al., 2020; Bobevski et al., 2016; Stults-Kolehmainen et al., 2014).

In this study, we found that there was a main effect of time for stress, anxiety, and depression, where the severity of each mental health condition became worse throughout time. Furthermore, all conditions increased significantly at each time interval. For example, depression at time 3 was significantly more severe than depression at time 2, which was more severe than depression at time 1; this effect was seen for anxiety and stress as well. We did not find a significant main effect of gender or perception of risk due to comorbidity on stress or depression. However, a main effect of risk perception was found on anxiety where individuals without a comorbidity that increased perception of lethality risk had lower anxiety than those who had a comorbidity. A time-by-gender interaction was also found where females had higher scores of anxiety at time 3 compared to males. Finally, a time-by-risk interaction was found where individuals with a comorbidity had lower anxiety scores than those without a comorbidity at time 3. The comorbidities that we asked participants can all be described as chronic illnesses, which include obesity, diabetes, high blood pressure and other cardiac and respiratory conditions (Pinto et al., 2020). We hypothesize that the reason that these individuals had lower anxiety scores is due to an increase in resiliency to stressful life events in this population. This hypothesis is consistent with previous literature, as Ghanei Gheshlagh et al. (2016) found that individuals with chronic illness have high resiliency scores, which increase with the disease lethality. These authors propose that high resiliency in these samples is developed adaptively, to maintain control of their own life, to adapt to life changes, and to remain in treatment, and for other reasons. Therefore, it is plausible that this sample of people with chronic illness/comorbidities was less anxious because of the COVID-19 pandemic as they had higher pre-existing resiliency than individuals without chronic illness.

We found that severity of all mental illness conditions was positively associated with each other at time one and negatively associated with age at time 1. At time 2, the only significant association was between age and depression. Subsequently, there were no significant associations between any of the conditions with each other or age at time 3. We hypothesize that the start of the pandemic provided a highly salient event that provoked similar difficulties across the sample. This universally shared experience may have led to mental health changes that were largely associated. However, as the pandemic progressed, individuals experienced different difficulties. For example, some may have experienced the loss of multiple loved ones, others may have lost employment, some may have endured the lockdown in solitude. Therefore, it is plausible that the conditions became dissociated as individuals developed unique concerns based on their distinct circumstance throughout the pandemic.

Our findings showed negative associations between age and mental health outcomes, which is consistent with other literature. More specifically, Huang and Zhao (2020) reported similar results; they found that younger Chinese presented higher scores in anxiety and depression. The same phenomenon occurred in both Germany (Bäuerle et al., 2020) and the U.S.A. (Liu et al., 2020) suggesting that, in fact, youth felt more impacted by the COVID-19 pandemic, which lead to worse mental health. Loneliness, anger and hopelessness were dimensions linked to depression and anxiety among young adults in U.S.A. (Liu et al., 2020) and among Brazilians (Goularte et al., 2021). Those symptoms affect younger individuals greater; thus, one might expect to observe a decrease of mental illness scores with increasing age.

We also collected data from non-binary gender participants (*N*=9). Even though only three of these volunteers answered our questionnaire at time1 and only one answered at time 2, it was possible to calculate an average and standard deviation for them at time 3, when most of them participated (*N*=8). At time 3, the average for perceived psychological stress in non-binary individuals was 26.3±12.4 versus 31.6±28.9 among females and 28.9±6.4 among males. Depression was 65.1±29.7 versus 81.7±30.0 and 69.5±22.5 among females and males, respectively. Lastly, anxiety was 59.4±17.5 among non-binary versus 66.7±11.8 and 57.4±13.5 among females and males, respectively. Even though it is not possible to perform a trustworthy null-hypothesis test due to the low number of non-binary participants, it seems that their scores were typically higher than other genders. The matter of mental health among binary and non-binary individuals is still a debatable, although researchers agree that cisgender (those whose gender is the same as the assigned at birth) tend to show better psychological outcomes when compared to transgender binary and non-binary participants (Rimes et al., 2019; Thorne et al., 2019). Nevertheless, it seems that stressful events, such as COVID-19 pandemic might be worse for those individuals. Specific studies should be conducted within this population to reach a more precise conclusion.

Limitations include the self-reported nature of the data, which was used to analyze mental health outcomes. Further, participants were asked if they had an illness which increased fatality risk for COVID-19 and were given a few examples of such conditions (i.e., obesity, diabetes, high blood pressure and other cardiac and respiratory conditions). Therefore, outside of these categories given to participants, the answer to this question relies on their knowledge of comorbidities that increase fatality risk for COVID-19 and their perception to the risk. For example, an individual with Vitamin D deficiency may be unaware of their status and state that they do not have any condition that increases risk for COVID-19 lethality, even though this condition increases fatality risk for COVID-19 (World Health Organization, 2017; Pugach and Pugach, 2021). Our study sheds light on changes in psychological stress, depression, and anxiety throughout early course of the COVID-19 pandemic curve in 2020. Prevalence, incidence, and severity increased for stress, anxiety, and depression. The largest increase in stress and anxiety incidence occurred between T2 and T3, while the largest increase in depression incidence occurred between T1 and T2. Further, our results suggests that as time progressed, scores of stress and depression significantly increased at similar rates in both genders and risk groups. However, anxiety scores increased at a higher rate in females and interestingly, individuals without a comorbidity that increased fatality risk. Additionally, we found a substantial decrease in the proportion of people that did not have a severe mental health condition.

## Data Availability

Data available upon reasonable request

